# Mechanical Dispersion Discriminates between Arrhythmic and Non-Arrhythmic Sudden Death: From the POST SCD Study

**DOI:** 10.1101/2023.05.22.23290353

**Authors:** Lionel Tastet, Satvik Ramakrishna, Lisa J. Lim, Dwight Bibby, Jeffrey E. Olgin, Andrew J. Connolly, Ellen Moffatt, Zian H. Tseng, Francesca N. Delling

## Abstract

**Background:** Global longitudinal strain (GLS) and mechanical dispersion (MD) by speckle-tracking echocardiography can predict sudden cardiac death (SCD) beyond left ventricular ejection fraction (LVEF) alone. However, prior studies have presumed cardiac cause from EMS records or death certificates rather than gold-standard autopsies.

**Objectives:** We sought to investigate whether abnormal GLS and MD, reflective of underlying myocardial fibrosis, are associated with autopsy-defined sudden arrhythmic death (SAD) in a comprehensive postmortem study.

**Methods:** We identified and autopsied all World Health Organization-defined (presumed) SCDs ages 18-90 via active surveillance of out of hospital deaths in the ongoing San Francisco POstmortem Systematic InvesTigation of Sudden Cardiac Death (POST SCD) Study to refine presumed SCDs to true cardiac causes. We retrieved all available pre-mortem echocardiograms and assessed LVEF, LV-GLS, and MD. The extent of LV myocardial fibrosis was assessed and quantified histologically.

**Results:** Of 652 autopsied subjects, 65 (10%) had echocardiograms available for primary review, obtained at a mean 1.5 years before SCD. Of these, 37 (56%) were SADs and 29 (44%) were non-SADs; fibrosis was quantified in 38 (58%). SADs were predominantly male, but had similar age, race, baseline comorbidities, and LVEF compared to non-SADs (all p>0.05). SADs had significantly reduced LV-GLS (median: –11.4% versus –18.5%, p=0.008) and increased MD (median: 14.8 ms versus 9.4 ms, p=0.006) compared to non-SADs. MD was associated with total LV fibrosis by linear regression in SADs (r=0.58, p=0.002).

**Conclusion:** In this countywide postmortem study of all sudden deaths, autopsy-confirmed arrhythmic deaths had significantly lower LV-GLS and increased MD than non-arrhythmic sudden deaths. Increased MD correlated with higher histologic levels of LV fibrosis in SADs. These findings suggest that increased MD, which is a surrogate for the extent of myocardial fibrosis, may improve risk stratification and specification for SAD beyond LVEF.

**PERSPECTIVES:** *Competency in medical knowledge:* Mechanical dispersion derived from speckle tracking echocardiography provides better discrimination between autopsy-defined arrhythmic vs non-arrhythmic sudden death than LVEF or LV-GLS. Histological ventricular fibrosis correlates with increased mechanical dispersion in SAD.

*Translational outlook:* Speckle tracking echocardiography parameters, in particular mechanical dispersion, may be considered as a non-invasive surrogate marker for myocardial fibrosis and risk stratification in SCD.

## INTRODUCTION

Sudden cardiac death (SCD) is a leading cause of premature death worldwide (1). In the United States, the age– and sex-adjusted incidence of SCD is 60 per 100,000 individuals, with an estimated number of premature deaths ranging from 180,000 to 450,000 each year (2,3). However, these estimates rely on medical or emergency medical services (EMS) records and death certificates, with inherent inaccuracies that may overestimate the actual incidence of SCD (4). Thus, the true burden, predictors, and underlying mechanisms of SCD, in particular sudden arrhythmic death (SAD) potentially rescuable with implantable cardioverter-defibrillators (ICDs), (5–8) remain elusive because only a minority of these out-of-hospital deaths are confirmed by autopsy. The POstmortem Systematic InvesTigation of Sudden Cardiac Death (POST SCD) study was the first prospective countywide study of all presumed SCDs and demonstrated that only half of SCDs defined by conventional criteria were true SADs (9,10). Thus nearly all prior studies of risk stratification of SCD are contaminated by the nearly half of cases due to causes not rescuable with an ICD (11).

The management and risk stratification for SCD remain highly dependent on the assessment of left ventricular ejection fraction (LVEF) (12). Although LVEF is a readily and ubiquitous imaging parameter providing a global measure of heart function, it has several limitations (load dependent and measurement variability), and may underestimate the extent of left ventricular (LV) systolic dysfunction (13). As a result, the risk predictive ability of LVEF alone has been challenged (14), suggesting that additional imaging diagnostic tools may provide better risk stratification for SCD. Speckle tracking echocardiography (STE) provides detailed assessment of LV global longitudinal strain (LV-GLS) and mechanical dispersion (MD), a measure of heterogeneous contraction of myocardial segments. Prior studies have shown that MD is linked to an increased risk of arrhythmic events in ischemic heart disease, dilated cardiomyopathy, and mitral valve prolapse (15–17), all conditions with underlying fibrosis independent of LVEF.

We sought to investigate the association between premortem LV-GLS, MD, and the extent of LV fibrosis in autopsy-defined arrhythmic versus non-arrhythmic sudden deaths in the POST SCD.

## METHODS

### Study population

The purpose and design of the POST SCD study have been previously described (18). Briefly, all incident presumed SCDs defined by World Health Organization (WHO) criteria in San Francisco County aged 18 to 90 years between February 2011 and May 2015 were prospectively identified for detailed autopsy, including toxicology and histology (9). SCD cases that met WHO criteria were fully adjudicated (9,18).

Out-of-hospital cardiac arrest (OHCA) deaths were defined as follows (18): ***i)*** deaths in the field or emergency department if the event was witnessed and/or active resuscitation was performed; or ***ii)*** unwitnessed natural deaths if the victim was last observed alive and symptom-free within 24 hours, with no active resuscitation, but with the impression of cardiac arrest by EMS. The following conditions were excluded: ***i)*** terminal illness; ***ii)*** end– stage renal disease on dialysis; ***iii)*** non-cardiac related deaths, including drug abuse/overdose or definite life-threatening trauma, homicide, or suicide; ***iv)*** hospital admission within the prior 30 days for non-cardiac conditions or surgery.

The POST SCD study was approved by the University of California San Francisco institutional review board, with additional review board of all 10 participating of adult hospitals and 3 EMS agencies serving the entire metropolitan area of San Francisco County.

### Postmortem investigation

After excision and dissection, internal organs of the thorax, abdomen, and cranial vault were examined to exclude non-cardiac etiologies. Vitreous chemistries were performed to rule out electrolyte abnormalities, and toxicology was performed on blood and urine. All hearts of presumed SCDs were examined for gross evidence of cardiovascular pathology and coronary artery disease as previously described (9).

Characterization of LV myocardial fibrosis was available in 38 of 65 (58%) cases. Histological 5 µm thick sections (including septum, posterobasal, lateral, and midanterior LV free wall) were stained with hematoxylin-eosin and Heidenhain trichrome and independently examined by 2 pathologists (E.M. and A.C.) (9,10). Total LV fibrosis (replacement + endoperimysial) was quantified using Aperio ImageScope as the average % fibrosis area of all four LV sections.

### Adjudication of sudden cardiac and arrhythmic deaths

Cause of death adjudication methods in the POST SCD Study have been previously described (9). Briefly, a multidisciplinary committee comprised of two electrophysiologists, the assistant medical examiner of San Francisco County, a cardiac pathologist, and a neurologist reviewed EMS and comprehensive medical records of all OHCA deaths to determine those that met WHO criteria for SCD. Autopsy-defined SAD was defined as those presumed SCDs potentially rescuable with ICD, without an obvious non-arrhythmic (e.g., tamponade, acute heart failure) or extra-cardiac (e.g., pulmonary embolism, occult overdose) cause of death. Resuscitated individuals who suffered OHCA and survived hospitalization were considered to be survivors of cardiac arrest rather than SCD because they did not die suddenly (19).

### Standard echocardiography

We identified victims of presumed SCD who underwent a transthoracic echocardiogram (TTE) as part of premortem routine clinical care and were available for review. Two-dimensional TTEs were performed using commercially available ultrasound systems. Left atrial (LA) volume, LV end-diastolic/end-systolic volumes, and LV mass were measured and indexed to body surface area according to the recommendations of the American Society of Echocardiography and European Association of Cardiovascular Imaging (20). LVEF was measured by the biplane Simpson method.

We also calculated LA function index (LAFI) which combines LA emptying fraction, indexed LA volume, and stroke volume (21).

### Speckle-tracking echocardiography

Speckle-tracking analysis was performed offline using a commercially available software (Tomtec Image Arena V.4.6) to measure LA and LV strain. For LA strain analysis, the LA endocardium was traced in both 4-chamber and 2-chamber views and the region of interest adjusted to the thinner wall of the atrium (22). LA strain analysis was calculated by averaging six LA segments from the apical 4-chamber and 2-chamber views (22). For LV strain analysis, the LV endocardium was traced at end-systole for each image in the 4-chamber, 3-chamber, and 2-chamber views. First, speckle-tracking of each of the 16 LV segments was conducted throughout the cardiac cycle. Then, the peak systolic strain was obtained for each segment and averaged for all segments as a measure of LV-GLS (17). The time from onset Q/R wave on electrocardiogram to the point of peak strain was defined as the time to peak strain. MD was defined as the standard deviation of the time to peak strain in 16 LV segments (15). The reproducibility of MD has been previously reported (17).

### Statistical analysis

Continuous variables were presented as mean ± standard deviation (SD) or median (interquartile range [IQR]) for non-normally distributed variables. Continuous variables with normal distribution were compared between groups with Student’s *t* test. Continuous variables non-normally distributed were compared with Wilcoxon-Mann-Whitney test. Categorical variables were presented as frequencies and percentages and were compared with *χ*^2^ test or Fisher’s exact test as appropriate. Relationship between STE parameters and myocardial fibrosis were determined using the Pearson or Spearman correlation coefficient, as appropriate.

Univariable and multivariable logistic regression analyses were performed to identify the clinical and imaging factors associated with SAD. The multivariable model included all clinically relevant variables and those significantly associated with SAD in univariable analysis (i.e. p<0.05). Results were presented as odds ratio (OR) with 95% confidence interval (CI). Multiple nested linear regression analyses were performed to identify the factors associated with the extent of myocardial fibrosis (i.e., total LV fibrosis). The variables included in the nested models were selected based on their relevant clinical value and/or significant association in univariable analysis (i.e. p<0.05). Results were presented as standardized beta (βeta) ± standard error (SE).

A two-tailed p value <0.05 was considered significant. Statistical analyses were performed with Stata software version 14.2 (StataCorp, College Station, Texas).

## RESULTS

### Study population

Of the 652 presumed SCDs who underwent a comprehensive autopsy and adjudication, 153 (24%) subjects had premortem TTEs available. Of these subjects, 100 had images available for review, but only 65 subjects had adequate image quality for 2D STE analysis (**Figure 1**). Mean time from last TTE to sudden death was 1.5±1.4 years. Among these, 36 (56%) were SADs and 29 (45%) non-SADs after adjudication. Compared to non-SADs, SADs were similar in age and prevalence of comorbidities including hypertension, smoking, coronary artery disease, congestive heart failure, valvular heart diseases, and atrial fibrillation but less likely to be women (19% versus 48%, p=0.01, **Table 1**). There was a trend toward more prescriptions for antiarrhythmics and calcium-channel blockers in non-SAD cases, while use of beta-blockers and anti-renin angiotensin system agents was comparable between groups (**Table 1**).

**FIGURE 1:**
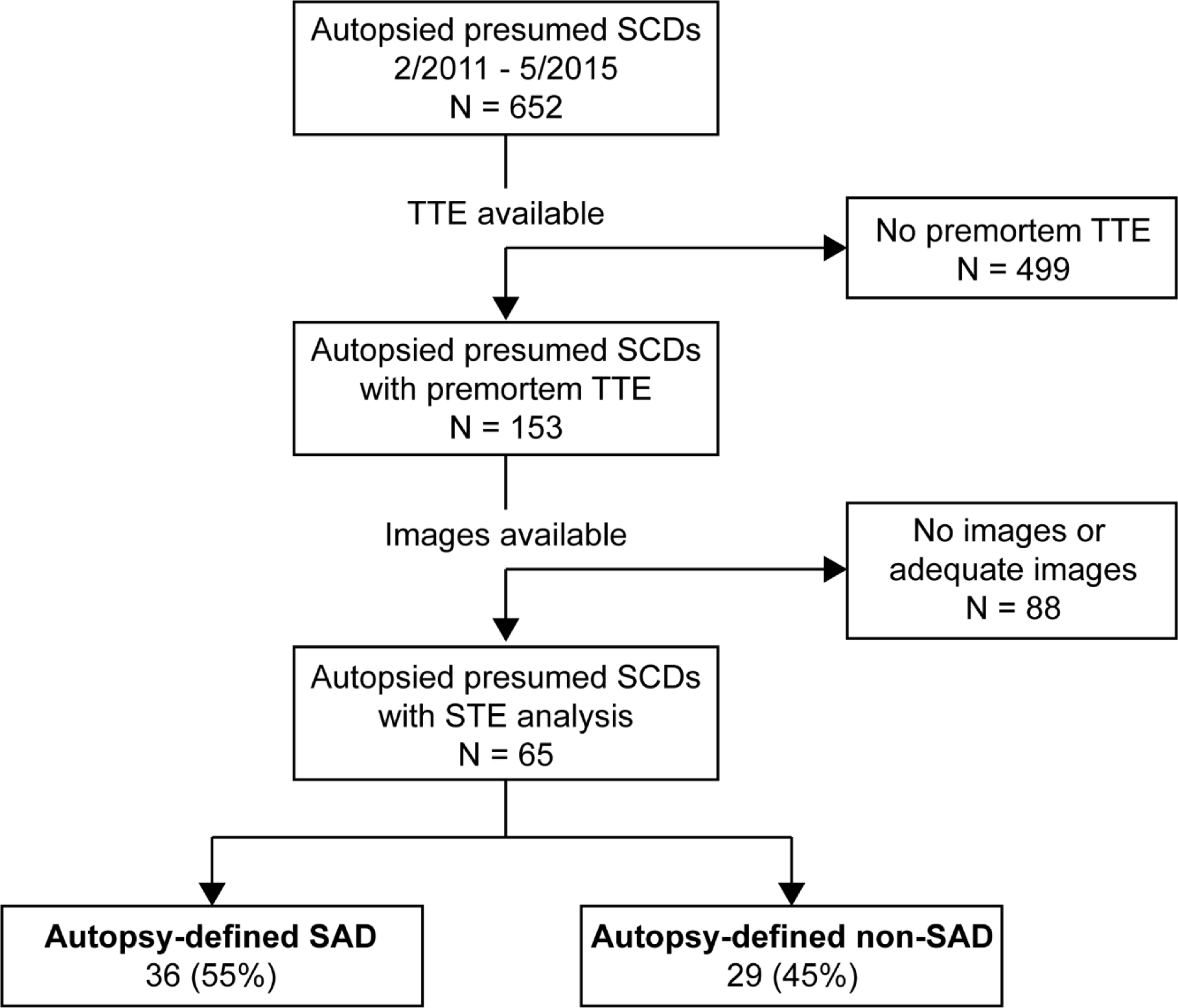
Study Flow Chart. LV = left ventricular; non-SAD = sudden non-arrhythmic death; SAD = sudden arrhythmic death; SCD = sudden cardiac death; TTE = transthoracic echocardiography.

**TABLE 1:**
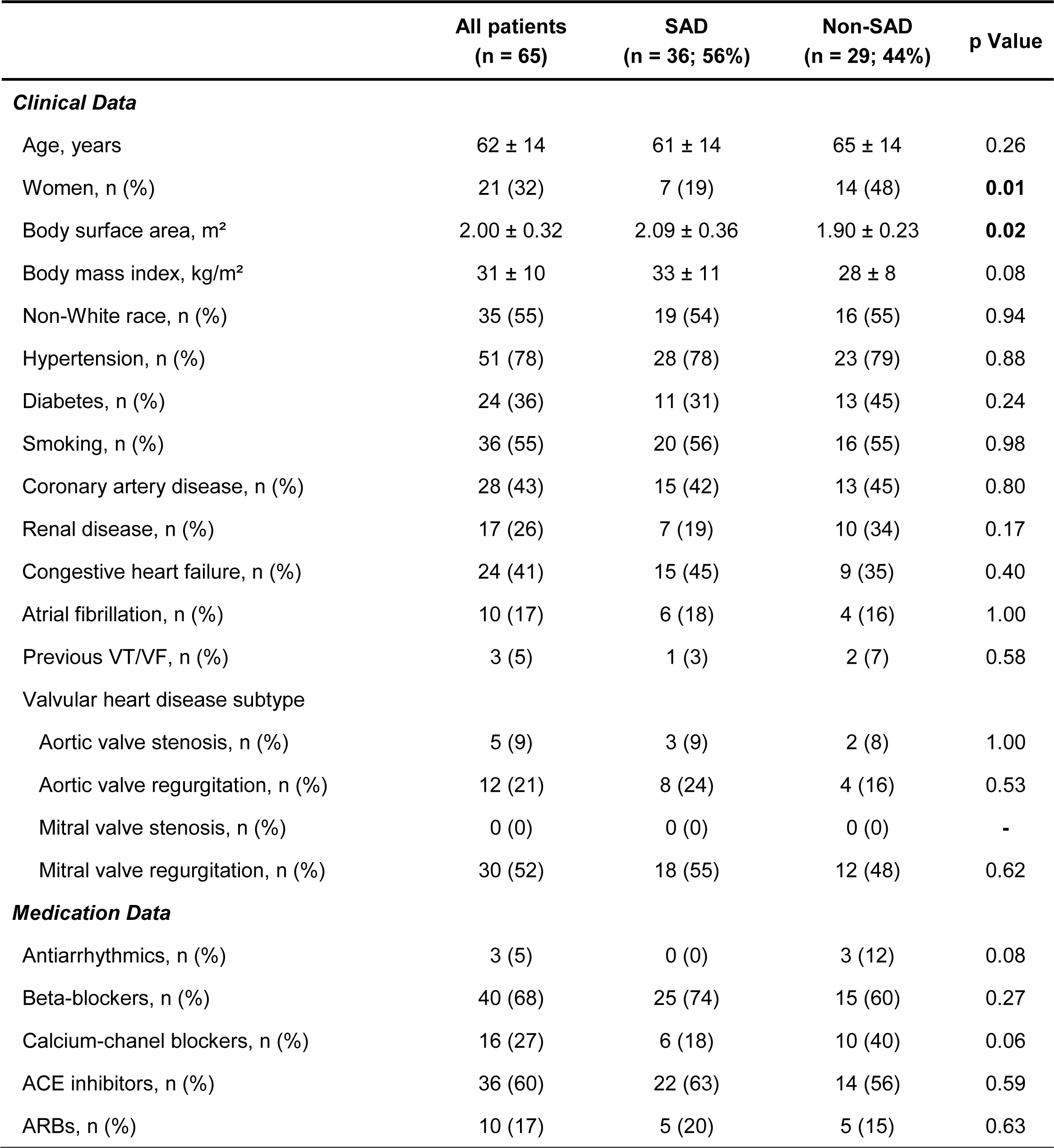
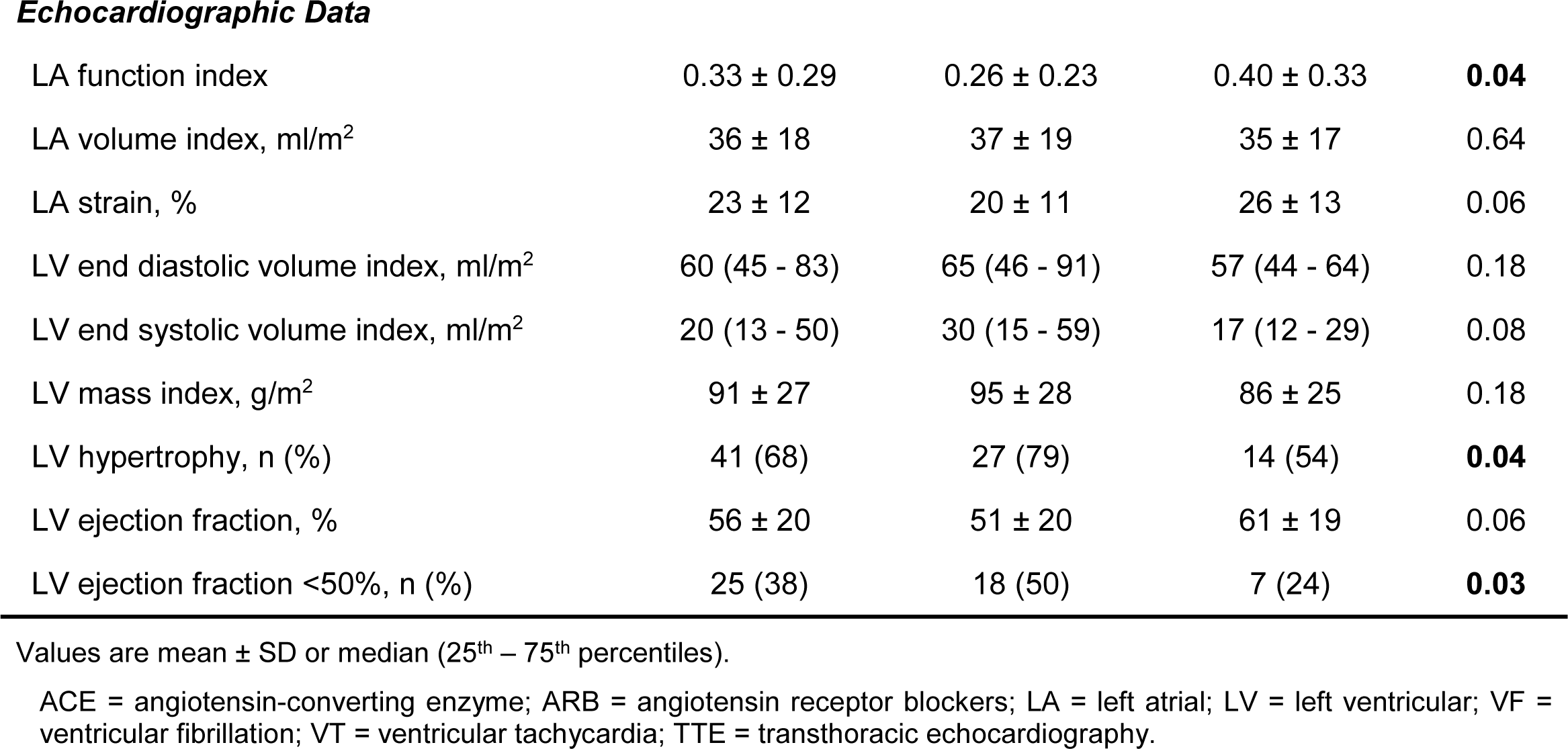
Premortem Characteristics of The Study Population with TTE.

|  | All patients<br>(n = 65) | SAD<br>(n = 36; 56%) | Non-SAD<br>(n = 29; 44%) | p Value |
| --- | --- | --- | --- | --- |
| **Clinical Data** |  |  |  |  |
| Age, years | 62 ± 14 | 61 ± 14 | 65 ± 14 | 0.26 |
| Women, n (%) | 21 (32) | 7 (19) | 14 (48) | **0.01** |
| Body surface area, m<sup>2</sup> | 2.00 ± 0.32 | 2.09 ± 0.36 | 1.90 ± 0.23 | **0.02** |
| Body mass index, kg/m<sup>2</sup> | 31 ± 10 | 33 ± 11 | 28 ± 8 | 0.08 |
| Non-White race, n (%) | 35 (55) | 19 (54) | 16 (55) | 0.94 |
| Hypertension, n (%) | 51 (78) | 28 (78) | 23 (79) | 0.88 |
| Diabetes, n (%) | 24 (36) | 11 (31) | 13 (45) | 0.24 |
| Smoking, n (%) | 36 (55) | 20 (56) | 16 (55) | 0.98 |
| Coronary artery disease, n (%) | 28 (43) | 15 (42) | 13 (45) | 0.80 |
| Renal disease, n (%) | 17 (26) | 7 (19) | 10 (34) | 0.17 |
| Congestive heart failure, n (%) | 24 (41) | 15 (45) | 9 (35) | 0.40 |
| Atrial fibrillation, n (%) | 10 (17) | 6 (18) | 4 (16) | 1.00 |
| Previous VT/VF, n (%) | 3 (5) | 1 (3) | 2 (7) | 0.58 |
| Valvular heart disease subtype |  |  |  |  |
| Aortic valve stenosis, n (%) | 5 (9) | 3 (9) | 2 (8) | 1.00 |
| Aortic valve regurgitation, n (%) | 12 (21) | 8 (24) | 4 (16) | 0.53 |
| Mitral valve stenosis, n (%) | 0 (0) | 0 (0) | 0 (0) | - |
| Mitral valve regurgitation, n (%) | 30 (52) | 18 (55) | 12 (48) | 0.62 |
| **Medication Data** |  |  |  |  |
| Antiarrhythmics, n (%) | 3 (5) | 0 (0) | 3 (12) | 0.08 |
| Beta-blockers, n (%) | 40 (68) | 25 (74) | 15 (60) | 0.27 |
| Calcium-channel blockers, n (%) | 16 (27) | 6 (18) | 10 (40) | 0.06 |
| ACE inhibitors, n (%) | 36 (60) | 22 (63) | 14 (56) | 0.59 |
| ARBs, n (%) | 10 (17) | 5 (20) | 5 (15) | 0.63 |
| ***Echocardiographic Data*** |  |  |  |  |
| LA function index | 0.33 ± 0.29 | 0.26 ± 0.23 | 0.40 ± 0.33 | **0.04** |
| LA volume index, ml/m<sup>2</sup> | 36 ± 18 | 37 ± 19 | 35 ± 17 | 0.64 |
| LA strain, % | 23 ± 12 | 20 ± 11 | 26 ± 13 | 0.06 |
| LV end diastolic volume index, ml/m<sup>2</sup> | 60 (45 - 83) | 65 (46 - 91) | 57 (44 - 64) | 0.18 |
| LV end systolic volume index, ml/m<sup>2</sup> | 20 (13 - 50) | 30 (15 - 59) | 17 (12 - 29) | 0.08 |
| LV mass index, g/m<sup>2</sup> | 91 ± 27 | 95 ± 28 | 86 ± 25 | 0.18 |
| LV hypertrophy, n (%) | 41 (68) | 27 (79) | 14 (54) | **0.04** |
| LV ejection fraction, % | 56 ± 20 | 51 ± 20 | 61 ± 19 | 0.06 |
| LV ejection fraction <50%, n (%) | 25 (38) | 18 (50) | 7 (24) | **0.03** |

Values are mean ± SD or median (25<sup>th</sup> – 75<sup>th</sup> percentiles).
ACE = angiotensin-converting enzyme; ARB = angiotensin receptor blockers; LA = left atrial; LV = left ventricular; VF = ventricular fibrillation; VT = ventricular tachycardia; TTE = transthoracic echocardiography.

### Standard echocardiography

Despite similar LA size between SAD and non-SAD (37±19 vs 35±17 ml/m^2^; p=0.64), SAD cases had lower LA function index (0.26±0.23 versus 0.40±0.33, p=0.04). SADs had similar LV dimensions (all p≤0.18), but more LV hypertrophy (p=0.04) compared to non-SADs (**Table 1**). Moreover, there was a trend toward a lower LVEF in SAD compared to non-SAD (p=0.06), although LVEF was normal in both groups (**Table 1**).

### Strain, mechanical dispersion and SAD

The **Central Illustration** shows the comparison of STE parameters between SAD and non-SAD. The median LV-GLS was significantly reduced in SADs versus non-SADs (– 11.4 % versus –18.5 %, p=0.008), while SADs cases showed significantly higher MD (14.8 ms versus 9.4 ms, p=0.006). We performed univariable and multivariable logistic regression analyses to identify the predictors of SAD (**Table 2**). Male sex, reduced LV-GLS and greater MD were significant predictors of SAD on univariate analysis (**Table 2**), but not LA strain or LAFI. Of note, we found a trend toward a significant association between lower LVEF and SAD (p=0.06, **Table 2**). On multivariable analysis adjusted for age, sex, and LVEF, MD remained the only significant predictor of SAD (OR: 1.69, 95% CI 1.06 – 2.68; p=0.03, **Table 2**). Including LV-GLS in the same multivariable model attenuated the association between MD and SAD (OR: 1.55, 95% CI 0.93 – 2.58; p=0.09) and LV-GLS and SAD (p=0.48) (**Table 2**).

**TABLE 2:** Factors Associated wtih Sudden Arrhythmic Death.

| Variables | Univariable Analysis |  | Multivariable Analysis #1 |  | Multivariable Analysis #2 |  |
| --- | --- | --- | --- | --- | --- | --- |
|  | OR (95% CI) | p Value | OR (95% CI) | p Value | OR (95% CI) | p Value |
| Age, per year increase | 0.98 (0.94 - 1.02) | 0.26 | 0.99 (0.94 - 1.03) | 0.49 | 0.99 (0.95 - 1.03) | 0.60 |
| Male sex | **3.87 (1.29 - 11.6)** | **0.02** | 2.93 (0.87 - 9.86) | 0.08 | 2.80 (0.82 - 9.51) | 0.10 |
| LA strain, per 1% decrease | 0.96 (0.92 - 1.00) | 0.06 | - | - | - | - |
| LA function index, per decrease | 0.13 (0.02 - 1.04) | 0.06 | - | - | - | - |
| LVEF, per 5% decrease | 0.88 (0.78 - 1.01) | 0.06 | 0.95 (0.82 - 1.10) | 0.49 | 0.99 (0.81 - 1.22) | 0.96 |
| LV-GLS, per 5% increase | **1.83 (1.16 - 2.89)** | **0.009** | - | - | 1.30 (0.62 - 2.75) | 0.48 |
| Mechanical dispersion, per 5 ms increase | **1.74 (1.11 - 2.71)** | **0.01** | **1.69 (1.06 - 2.68)** | **0.03** | 1.55 (0.93 - 2.58) | 0.09 |
| Myocardial fibrosis, per 1% increase | 1.01 (0.94 - 1.09) | 0.76 | - | - | - | - |

Results are presented as odd ratio (OR) with 95% confidence interval (CI). Bold text indicates significant association (p<0.05).
LVEF = left ventricular ejection fraction; LV-GLS = left ventricular global longitudinal strain; other abbreviations as in Table 1.

### Mechanical dispersion and myocardial fibrosis

Among the 65 presumed SCD cases with echocardiographic strain assessment, 38 (58%) had characterization of burden and type of myocardial fibrosis. Replacement fibrosis was detected in 24 (63%) of 38 cases, 25 (69%) of which were SADs and 13 (45%) non-SADs (**Figure 1**).

We found a significant correlation between MD and total LV myocardial fibrosis in the entire sample (**Figure 2A** and **Central Illustration**). MD was associated with the extent of LV myocardial fibrosis in the SADs (p=0.002), but not non-SADs (p=0.20, **Figures 2B** and **2C**). Of note, LV-GLS was not associated with fibrosis in the entire sample of presumed SCDs, nor in SADs versus non-SADs (all p≥0.10).

**FIGURE 2:**
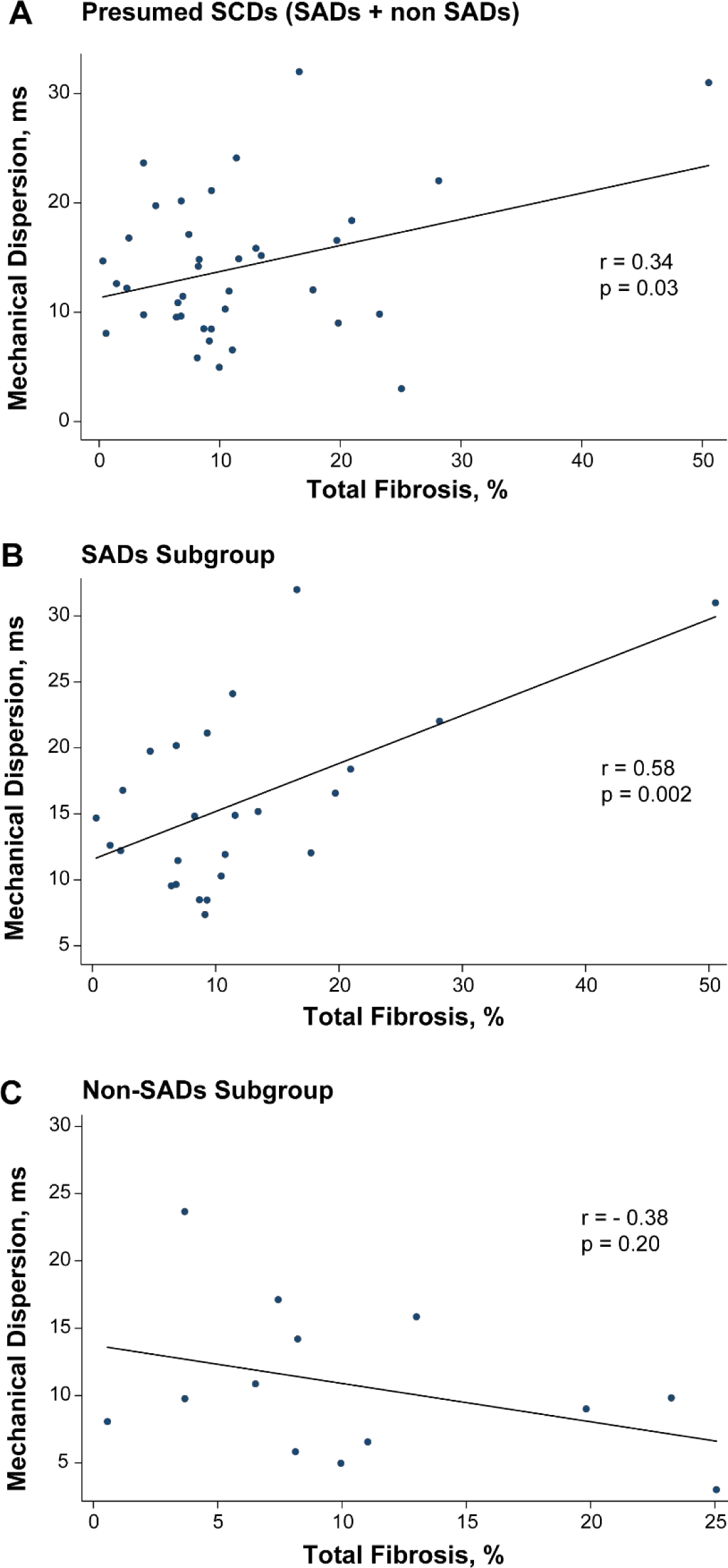
Relationship Between Myocardial Fibrosis and Mechanical Dispersion. ***Caption***: Correlation between total myocardial fibrosis histologically quantified and mechanical dispersion in the whole study population (n=38) **(A)**, and according to sudden arrhythmic death (SAD) **(B)** and non-SAD **(C)**.

To identify the independent predictors of myocardial fibrosis among SADs (n=25), we performed several nested regression models inclusive of clinical and/or imaging factors (**Table 3**). In all models, MD remained a significant predictor of LV myocardial fibrosis among SADs (all p≤0.02) (**Table 3**). MD remained significantly associated with myocardial fibrosis after adjustment for LVEF and LV-GLS (p=0.001, **Table 3**).

**TABLE 3:**
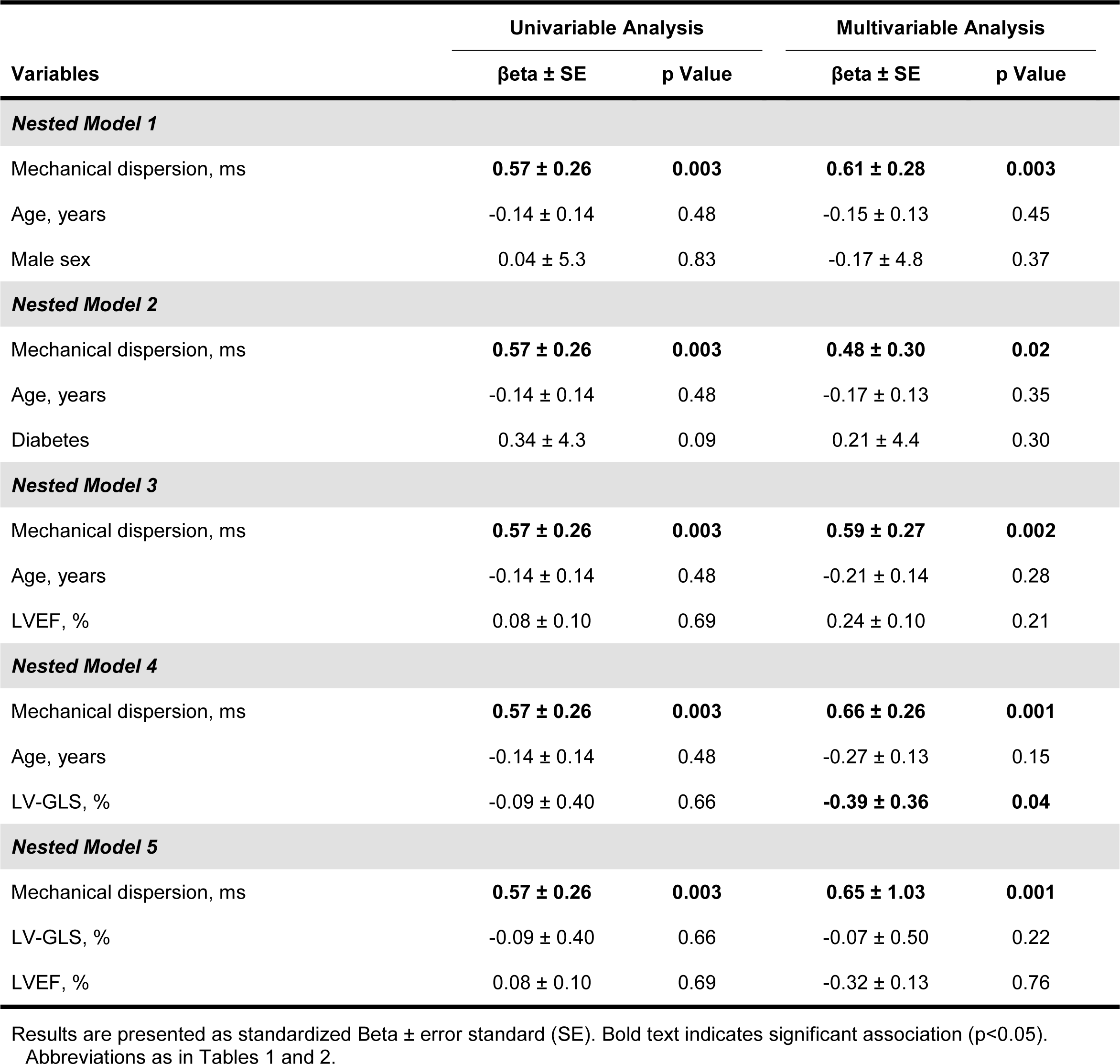
Predictors of Myocardial Fibrosis in the Sudden Arrhythmic Death Subgroup.

| Variables | Univariable Analysis |  | Multivariable Analysis |  |
| --- | --- | --- | --- | --- |
| | $\beta$ ± SE | p Value | $\beta$ ± SE | p Value |
| ***Nested Model 1*** |  |  |  |  |
| Mechanical dispersion, ms | **0.57 ± 0.26** | **0.003** | **0.61 ± 0.28** | **0.003** |
| Age, years | -0.14 ± 0.14 | 0.48 | -0.15 ± 0.13 | 0.45 |
| Male sex | 0.04 ± 5.3 | 0.83 | -0.17 ± 4.8 | 0.37 |
| ***Nested Model 2*** |  |  |  |  |
| Mechanical dispersion, ms | **0.57 ± 0.26** | **0.003** | **0.48 ± 0.30** | **0.02** |
| Age, years | -0.14 ± 0.14 | 0.48 | -0.17 ± 0.13 | 0.35 |
| Diabetes | 0.34 ± 4.3 | 0.09 | 0.21 ± 4.4 | 0.30 |
| ***Nested Model 3*** |  |  |  |  |
| Mechanical dispersion, ms | **0.57 ± 0.26** | **0.003** | **0.59 ± 0.27** | **0.002** |
| Age, years | -0.14 ± 0.14 | 0.48 | -0.21 ± 0.14 | 0.28 |
| LVEF, % | 0.08 ± 0.10 | 0.69 | 0.24 ± 0.10 | 0.21 |
| ***Nested Model 4*** |  |  |  |  |
| Mechanical dispersion, ms | **0.57 ± 0.26** | **0.003** | **0.66 ± 0.26** | **0.001** |
| Age, years | -0.14 ± 0.14 | 0.48 | -0.27 ± 0.13 | 0.15 |
| LV-GLS, % | -0.09 ± 0.40 | 0.66 | **-0.39 ± 0.36** | **0.04** |
| ***Nested Model 5*** |  |  |  |  |
| Mechanical dispersion, ms | **0.57 ± 0.26** | **0.003** | **0.65 ± 1.03** | **0.001** |
| LV-GLS, % | -0.09 ± 0.40 | 0.66 | -0.07 ± 0.50 | 0.22 |
| LVEF, % | 0.08 ± 0.10 | 0.69 | -0.32 ± 0.13 | 0.76 |

Results are presented as standardized Beta ± error standard (SE). Bold text indicates significant association (p<0.05). Abbreviations as in Tables 1 and 2.

We then compared the distribution of SADs versus non-SADs by median values of LV-GLS (i.e., –16%) and MD (i.e., 12 ms) and the presence of replacement fibrosis in the entire sample. Specifically, we assessed the number of SAD vs non-SADs with reduced LV-GLS (i.e., > –16%), increased MD (i.e., > 12 ms) or with replacement fibrosis (found in 24 cases in total, **Figure 3**). Of all SCDs with both reduced LV-GLS and increased MD, 77% were SAD vs. 23% non-SAD (**Figure 3**). This difference was greater among the cases meeting all criteria, i.e., reduced LV-GLS, high MD, and presence of replacement fibrosis (86% of SADs versus 13% of non-SADs, **Figure 3**).

**FIGURE 3:**
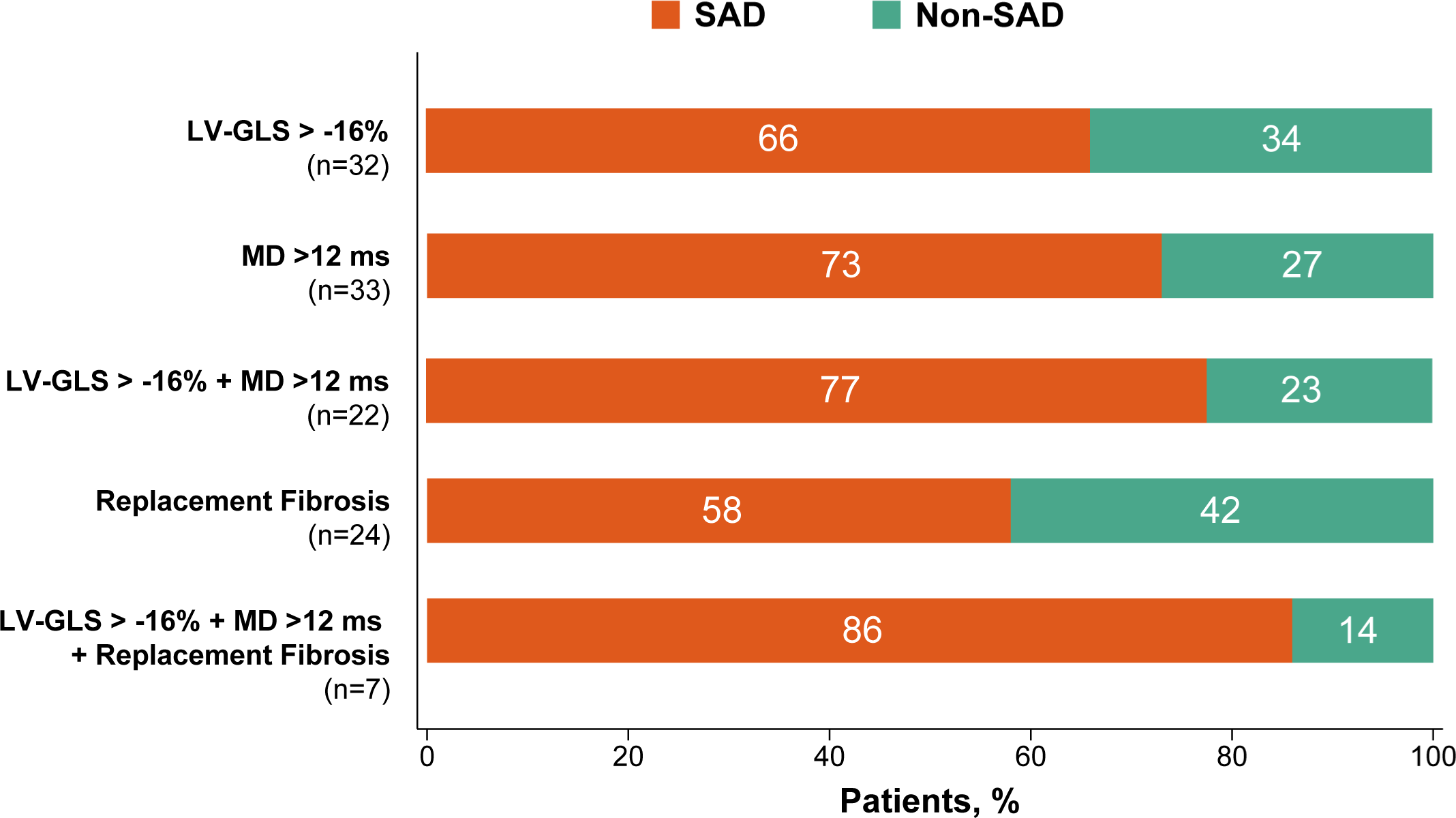
Distribution of Sudden Arrhythmic and Non-Arrhythmic Deaths According Speckle Tracking Echocardiography Parameters, and Myocardial Fibrosis. ***Caption***: Comparison of the distribution of sudden arrhythmic death (SAD) versus non-SAD according to the median values of LV-GLS (i.e. LV-GLS > –16%) and mechanical dispersion (i.e. >12 ms) in the entire sample, and the presence of replacement fibrosis. LV-GLS = left ventricular global longitudinal strain; MD = mechanical dispersion; other abbreviation as in Figure 1.

## DISCUSSION

In this countywide postmortem study of presumed SCDs, we found that: ***i)*** autopsy-confirmed arrhythmic death is associated with reduced LV-GLS and increased MD compared to non-arrhythmic causes of sudden death (**Central Illustration**); ***ii)*** MD provides better discrimination between arrhythmic and non-arrhythmic causes among sudden deaths compared to LVEF or LV-GLS. ***iii)*** the extent of myocardial fibrosis is strongly correlated with MD in SAD, but not in non-SADs.

**CENTRAL ILLUSTRATION:**
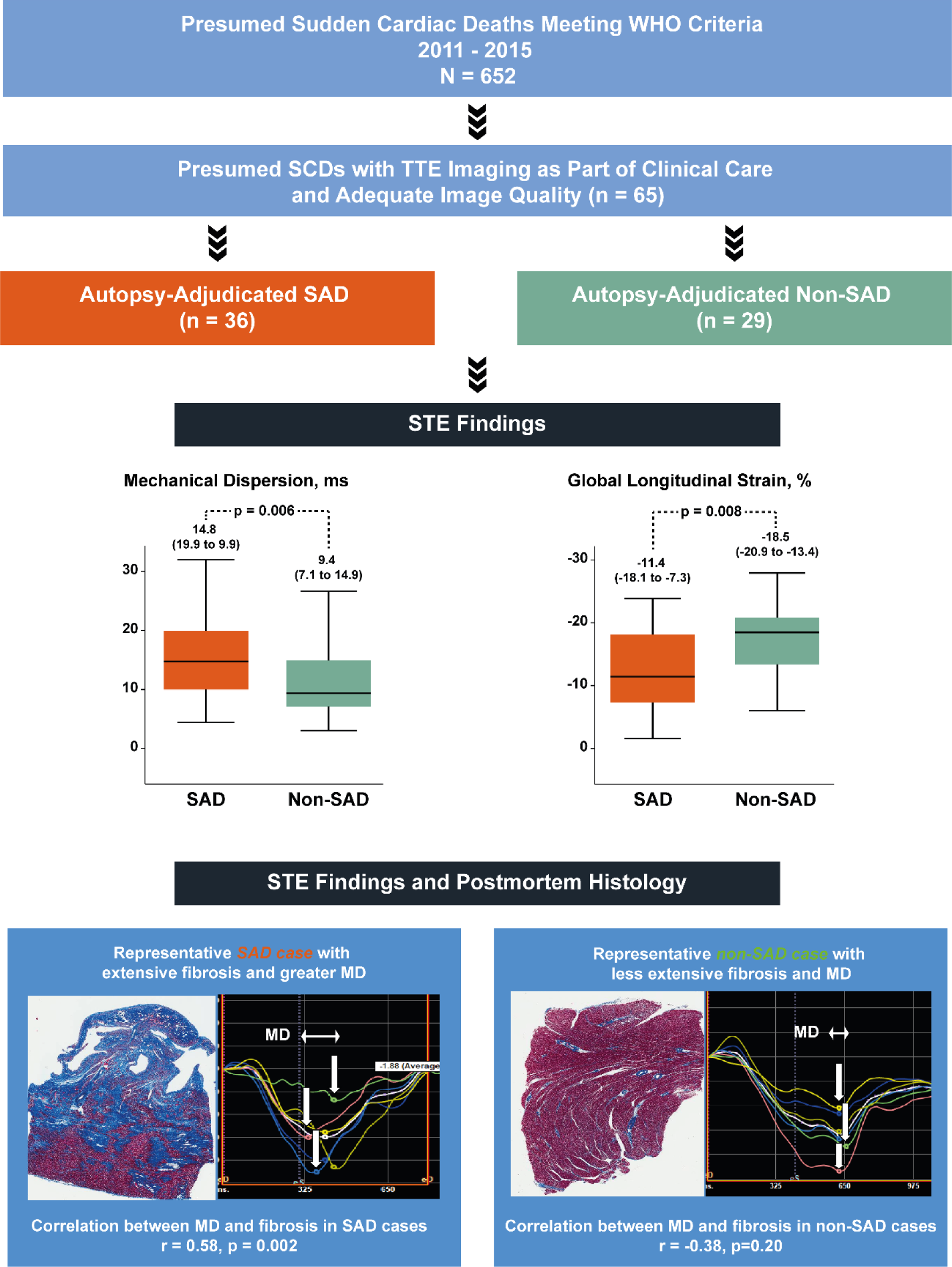
Speckle Tracking Echocardiography, Histological Myocardial Fibrosis, and Sudden Arrhythmic Death. ***Caption*:** Sudden arrhythmic death (SAD) cases had significantly greater mechanical dispersion and lower global longitudinal strain compared to non-SAD cases (**Top Figure**). Medians and 25th to 75th percentiles are shown on top of each box-plot. In 38 sudden cardiac death (SCD) cases with comprehensive histological analysis for myocardial tissue characterization, we observed a significant correlation between histological ventricular fibrosis and greater mechanical dispersion in SAD cases (r=0.58, p=0.002), but not in non-SAD cases (r=-0.38, p=0.20). This finding is illustrated by a representative case of SAD with more extensive fibrosis (blue stain by hematoxylin-eosin) and greater mechanical dispersion compared to a representative case of non-SAD (**Bottom Figure**). STE = speckle tracking echocardiography; other abbreviations as in Figures 1 and 3.

### Risk stratification of SCD

In recent years, the assessment of MD by STE has emerged as a useful imaging parameter to predict arrhythmic-related events in various cardiovascular settings (15-17, 23-25). Indeed, increased MD has been associated with recurrent arrhythmias after myocardial infarction (15). In patients with nonischemic dilated cardiomyopathy, MD and LV-GLS were more accurate than LVEF alone in predicting arrhythmic events (16). Moreover, reduced LV-GLS and abnormal MD were associated with genotyped long QT syndrome (25). Likewise, in patients with mitral valve prolapse, we also reported the significant and independent predictive value of MD for the risk of ventricular arrhythmias (17). To our knowledge, our study represents the first histological validation of STE parameters, including MD, in adjudicated cases of SCD after comprehensive autopsy examination. Indeed, a major limitation of prior studies assessing the performance of STE in predicting ventricular arrhythmias or SCD in cardiac conditions with associated myocardial fibrosis, was the lack of autopsy confirmation of cause of death, raising uncertainty about the specificity of arrhythmic death and the prognostic role of STE.

We investigate the performance of LV-GLS and MD in predicting true arrhythmic death by leveraging the POST SCD Study, a countywide postmortem investigation of all consecutive cases of SCD with detailed adjudication of cause of death (9). Based on this unbiased and precise approach, the present study demonstrates that SADs are associated with significantly reduced LV-GLS and increased MD compared to non-SADs cases, and may therefore better specify patients at risk for sudden death rescuable with ICD. We found that about 80% of SCDs with both LV-GLS and MD that exceeded the median value of this population were SADs. This observation further supports the usefulness of STE for prediction of arrhythmic events. Importantly, MD was associated with SAD risk independently of LVEF and other conventional risk factors. This is consistent with a recent meta-analysis of Haugaa et al. (26), where LVEF showed mediocre performance in predicting risk of ventricular arrhythmias compared to MD. In this regard, previous reports show that 30% to 40% of SCDs cases are occurring in individuals with mid-range to normal LVEF (i.e. LVEF >35%). In our study, over three-quarters of postmortem-confirmed cases had LVEF >35%, suggesting that the current guideline-directed criteria for SCA/SCD risk prevention are inadequately sensitive to identify a substantial proportion of high-risk patients. Furthermore, the predictive value of MD may also be superior to LV-GLS. Our results suggest that even in the presence of “a priori” normal LV function as assessed by LVEF or LV-GLS alone, some individuals with abnormal MD are still at increased risk of arrhythmic events. In fact, besides the ability to measure contractile heterogeneity of the LV, MD may represent a marker of electrophysiological heterogeneity, which also contributes to the occurrence of arrhythmic events (26). For example, longitudinal MD showed better risk discrimination than corrected QT for arrhythmia risk assessment in long QT syndrome (27). Hence, abnormal MD as detected by STE may also reflect electrical dispersion of repolarization (27). However, further experimental and clinical studies are needed to test this hypothesis.

In our study, SADs demonstrated worse LA function with lower LAFI and LA strain compared to non-SADs at baseline. Abnormal atrial function in SAD cases could be related to some extent to the greater proportion of cases with reduced ejection fraction, which is typically associated with increased LA pressure. Although atrial fibrillation and reduced LVEF frequently coexist, the prevalence of atrial fibrillation and LA size were similar between SAD cases and non-SAD cases. Whereas LV-GLS and MD were associated with SAD in regression analyses, LA strain and LAFI were not, supporting a stronger role of LV compared to LA parameters in SAD risk stratification in our sample.

### Increased mechanical dispersion, a surrogate marker of fibrosis

Despite numerous efforts over the past decades, the underlying pathophysiological mechanisms leading to SCD have not been completely unraveled. Myocardial fibrosis, in particular replacement fibrosis, has been proposed as a substrate for reentry (28,29), leading to ventricular arrhythmias and SCD (30) and is higher in systemic conditions such as HIV (10). Interestingly, increased MD, a marker of electrical dispersion, has been correlated to replacement fibrosis as detected by late gadolinium enhancement using cardiac magnetic resonance in patients with hypertrophic cardiomyopathy (31). However, no previous studies have examined whether the risk of fatal arrhythmias predicted by increased MD may be mediated by myocardial fibrosis as confirmed histologically. In the present study including autopsy-confirmed cases of SCD, we observed a significant association between LV fibrosis and MD, suggesting that MD measured by STE may be considered as a surrogate marker for myocardial fibrosis. In addition, we found a significant association between MD and myocardial fibrosis only among SADs. These findings support the hypothesis that the risk of fatal arrhythmias associated with increased MD is predominantly mediated by the extent of myocardial fibrosis. Further studies are needed to elucidate the underlying molecular mechanisms underlying the lethal arrhythmic risk associated with increased MD.

### Clinical Implications

Risk prediction and prevention of SCD remain a major clinical conundrum. Hence, the elaboration of a novel integrative approach that goes beyond LVEF alone, is essential to improve risk stratification in SCD. STE is a robust, accurate and readily accessible imaging modality in clinical practice. This study shows that STE-derived MD is a useful non-invasive imaging tool which correlates with histological myocardial fibrosis. Importantly, a substantial proportion of individuals in our postmortem sample who ultimately experienced SAD did not meet current, LVEF-focused guidelines. In contrast, our data suggests that individuals with LVEF ≥35% but abnormal MD may benefit from closer comprehensive clinical and imaging monitoring. Moreover, randomized clinical trials may be considered to address the additional value of STE for primary and/or secondary prevention of arrhythmic events.

### Study limitations

The relatively small sample size of our study limited the multivariable analyses as well as the identification of an optimal threshold to define abnormal MD, and reduced power to detect differences. However, these findings reflect the real-world population of victims of sudden death. We also showed in different nested multivariable models that MD was a strong and independent predictor of SAD. In addition, we demonstrate for the first the relationship between STE-derived parameters and myocardial fibrosis in SAD after comprehensive autopsy, toxicology, and histology. The time between TTE and sudden death was 1.5 years, however, these parameters may have actually increased had TTEs been performed more proximate to time of sudden death.

## CONCLUSIONS

In this countywide postmortem study of all presumed SCDs with premortem imaging obtained as part of typical clinical care, most SADs had normal LVEF, and increased MD discriminated between arrhythmic deaths potentially rescuable with ICD and non-arrhythmic causes of sudden death. Moreover, MD was strongly associated with the extent of LV myocardial fibrosis in SAD. These findings suggest that beyond LVEF, abnormal LV strain and increased MD, as surrogates for the extent of myocardial fibrosis, may significantly improve SAD risk stratification and discrimination.

## Data Availability

All data produced in the present study are available upon reasonable request to the authors.

## ABBREVIATIONS

SCD: sudden cardiac death
SAD: sudden arrhythmic death
EMS: emergency medical services
STE: speckle tracking echocardiography
LVEF: left ventricular ejection fraction
LV: left ventricular/ventricle
LV-GLS: left ventricular global longitudinal strain
MD: mechanical dispersion
OHCA: Out-of-hospital cardiac arrest
TTE: transthoracic echocardiogram
LA: left atrial
OR: odds ratio

## ACKNOWLEDGMENTS

We thank all victims of sudden death and their families in San Francisco County; Shiktij Dave and Justin Cheng for their assistance with data collection; and all forensic investigators in the Office of the Chief Medical Examiner and Emergency Medical Services personnel in San Francisco County.

## REFERENCES

1. Berdowski J, Berg RA, Tijssen JG, Koster RW. Global incidences of out-of-hospital cardiac arrest and survival rates: Systematic review of 67 prospective studies. Resuscitation 2010;81:1479–87.

2. Kong MH, Fonarow GC, Peterson ED et al. Systematic review of the incidence of sudden cardiac death in the United States. J Am Coll Cardiol 2011;57:794–801.

3. Stecker EC, Reinier K, Marijon E et al. Public health burden of sudden cardiac death in the United States. Circ Arrhythm Electrophysiol 2014;7:212–7.

4. Chugh SS, Jui J, Gunson K et al. Current burden of sudden cardiac death: multiple source surveillance versus retrospective death certificate-based review in a large U.S. community. J Am Coll Cardiol 2004;44:1268–75.

5. Bardy GH, Lee KL, Mark DB et al. Amiodarone or an implantable cardioverter-defibrillator for congestive heart failure. N Engl J Med 2005;352:225–37.

6. Buxton AE, Lee KL, Fisher JD, Josephson ME, Prystowsky EN, Hafley G. A randomized study of the prevention of sudden death in patients with coronary artery disease. Multicenter Unsustained Tachycardia Trial Investigators. N Engl J Med 1999;341:1882–90.

7. Moss AJ, Hall WJ, Cannom DS et al. Improved survival with an implanted defibrillator in patients with coronary disease at high risk for ventricular arrhythmia. Multicenter Automatic Defibrillator Implantation Trial Investigators. N Engl J Med 1996;335:1933–40.

8. Moss AJ, Zareba W, Hall WJ et al. Prophylactic implantation of a defibrillator in patients with myocardial infarction and reduced ejection fraction. N Engl J Med 2002;346:877–83.

9. Tseng ZH, Olgin JE, Vittinghoff E et al. Prospective Countywide Surveillance and Autopsy Characterization of Sudden Cardiac Death: POST SCD Study. Circulation 2018;137:2689–2700.

10. Tseng ZH, Moffatt E, Kim A, et al. Sudden Cardiac Death and Myocardial Fibrosis, Determined by Autopsy, in Persons with HIV. N Engl J Med 2021;384:2306–2316.

11. Benjamin EJ, Virani SS, Callaway CW, et al. Heart disease and stroke statistics – 2018 update: A report from the American Heart Association. Circulation 2018;137:e67–e492.

12. Al-Khatib SM, Stevenson WG, Ackerman MJ et al. 2017 AHA/ACC/HRS Guideline for Management of Patients With Ventricular Arrhythmias and the Prevention of Sudden Cardiac Death: A Report of the American College of Cardiology/American Heart Association Task Force on Clinical Practice Guidelines and the Heart Rhythm Society. J Am Coll Cardiol 2018;72:e91–e220.

13. Wellens HJ, Schwartz PJ, Lindemans FW et al. Risk stratification for sudden cardiac death: current status and challenges for the future. European heart journal 2014;35:1642–51.

14. Deyell MW, Krahn AD, Goldberger JJ. Sudden cardiac death risk stratification. Circ Res 2015;116:1907–18.

15. Haugaa KH, Smedsrud MK, Steen T et al. Mechanical dispersion assessed by myocardial strain in patients after myocardial infarction for risk prediction of ventricular arrhythmia. JACC Cardiovascular imaging 2010;3:247–56.

16. Haugaa KH, Goebel B, Dahlslett T et al. Risk assessment of ventricular arrhythmias in patients with nonischemic dilated cardiomyopathy by strain echocardiography. J Am Soc Echocardiogr 2012;25:667–73.

17. Ermakov S, Gulhar R, Lim L et al. Left ventricular mechanical dispersion predicts arrhythmic risk in mitral valve prolapse. Heart (British Cardiac Society) 2019;105:1063–1069.

18. Delling FN, Aung S, Vittinghoff E et al. Antemortem and Post-Mortem Characteristics of Lethal Mitral Valve Prolapse Among All Countywide Sudden Deaths. JACC Clin Electrophysiol 2021;7:1025–1034.

19. Ricceri S, Salazar JW, Vu AA, Vittinghoff E, Moffatt E, Tseng ZH. Factors Predisposing to Survival After Resuscitation for Sudden Cardiac Arrest. J Am Coll Cardiol 2021;77:2353–2362.

20. Lang RM, Badano LP, Mor-Avi V et al. Recommendations for cardiac chamber quantification by echocardiography in adults: an update from the american society of echocardiography and the European association of cardiovascular imaging. J Am Soc Echocardiogr 2015;28:1–39.

21. Thomas L, Hoy M, Byth K, Schiller NB. The left atrial function index: a rhythm independent marker of atrial function. Eur J Echocardiogr 2008;9:356–62.

22. Badano LP, Kolias TJ, Muraru D et al. Standardization of left atrial, right ventricular, and right atrial deformation imaging using two-dimensional speckle tracking echocardiography: a consensus document of the EACVI/ASE/Industry Task Force to standardize deformation imaging. European heart journal cardiovascular Imaging 2018;19:591–600.

23. Ersbøll M, Valeur N, Andersen MJ et al. Early echocardiographic deformation analysis for the prediction of sudden cardiac death and life-threatening arrhythmias after myocardial infarction. JACC Cardiovascular imaging 2013;6:851–60.

24. Haugaa KH, Grenne BL, Eek CH et al. Strain echocardiography improves risk prediction of ventricular arrhythmias after myocardial infarction. JACC Cardiovascular imaging 2013;6:841–50.

25. Leren IS, Hasselberg NE, Saberniak J et al. Cardiac mechanical alterations and genotype specific differences in subjects with long QT syndrome. JACC Cardiovascular imaging 2015;8:501–510.

26. Kawakami H, Nerlekar N, Haugaa KH, Edvardsen T, Marwick TH. Prediction of Ventricular Arrhythmias With Left Ventricular Mechanical Dispersion: A Systematic Review and Meta-Analysis. JACC Cardiovascular imaging 2020;13:562–572.

27. Haugaa KH, Amlie JP, Berge KE, Leren TP, Smiseth OA, Edvardsen T. Transmural differences in myocardial contraction in long-QT syndrome: mechanical consequences of ion channel dysfunction. Circulation 2010;122:1355–63.

28. Zhang J, Cooper DH, Desouza KA et al. Electrophysiologic Scar Substrate in Relation to VT: Noninvasive High-Resolution Mapping and Risk Assessment with ECGI. Pacing Clin Electrophysiol 2016;39:781–91.

29. Fernández-Armenta J, Penela D, Acosta J et al. Substrate modification or ventricular tachycardia induction, mapping, and ablation as the first step? A randomized study. Heart Rhythm 2016;13:1589–95.

30. Hayashi M, Shimizu W, Albert CM. The spectrum of epidemiology underlying sudden cardiac death. Circ Res 2015;116:1887–906.

31. Haland TF, Almaas VM, Hasselberg NE et al. Strain echocardiography is related to fibrosis and ventricular arrhythmias in hypertrophic cardiomyopathy. European Heart Journal – Cardiovascular Imaging 2016;17:613–621.

